# Pre-activated anti-viral innate immunity in the upper airways controls early SARS-CoV-2 infection in children

**DOI:** 10.1101/2021.06.24.21259087

**Authors:** J Loske, J Röhmel, S Lukassen, S Stricker, VG Magalhães, J Liebig, RL Chua, L Thürmann, M Messingschlager, A Seegebarth, B Timmermann, S Klages, M Ralser, B Sawitzki, LE Sander, VM Corman, C Conrad, S Laudi, M Binder, S Trump, R Eils, M.A. Mall, I Lehmann

## Abstract

Children are consistently reported to have reduced SARS-CoV-2 infection rates and a substantially lower risk for developing severe COVID-19. However, the molecular mechanisms underlying protection against COVID-19 in younger age groups remain widely unknown. Here, we systematically characterized the single-cell transcriptional landscape in the upper airways in SARS-CoV-2 negative and age-matched SARS-CoV-2 positive children (n=42) and corresponding samples from adults (n=44), covering an age range of four weeks to 77 years. Children displayed higher basal expression of the relevant pattern recognition receptor (PRR) pathways in upper airway epithelial cells, macrophages, and dendritic cells, resulting in stronger innate antiviral responses upon SARS-CoV-2 infection compared to adults. We further detected distinct immune cell subpopulations with an overall dominance of neutrophils and a population of cytotoxic T cells occurring predominantly in children. Our study provides evidence that the airway epithelial and mucosal immune cells of children are pre-activated and primed for virus sensing, resulting in a stronger early innate antiviral responses to SARS-CoV-2 infection compared to adults.

---

It has repeatedly been reported that younger individuals have a substantially lower risk for developing COVID-19, despite a similar risk of infection, as reflected in dramatically increased mortality with increasing age ^1-3^. These observations suggest that children may have a higher capability of controlling SARS-CoV-2 infection. It has been shown that an early cell-intrinsic innate immune response, mediated by pattern recognition receptors (PRR) and the type I and III interferon (IFN) system, are crucial for the successful control of SARS-CoV-2 infection ^4^. In line with these observations, recent studies compared adults and children with severe COVID-19 or those presenting to an Emergency Department and described an impaired IFN response in pediatric COVID-19 ^5, 6^. However, the molecular mechanisms protecting against COVID-19 in younger age groups particularly in those with no or only mild/moderate symptoms remain unknown.

To understand the higher capacity of children for controlling SARS-CoV-2 infection at an early stage we systematically characterized the transcriptional landscape of upper airways, an airway region with high susceptibility for SARS-CoV-2 infection ^7^, in SARS-CoV-2 negative and SARS-CoV-2 positive children (n=42) and adults (n=44), comprising 268,745 cells in total (Fig. 1a). To this end, we included study participants of three different COVID-19 cohorts: the RECAST study focusing on COVID-19 in children and their families, the Pa-COVID-19 and the SC2 study ^8, 9^. Samples from the upper airways (nose) were collected from individuals aged 4 weeks to 77 years with a positive SARS-CoV-2 PCR result along with age-matched SARS-CoV-2 negative controls (Suppl. Table 1, 2). Focusing on early infection only mild/moderate COVID-19 cases were considered for this study (Fig. 1a). Based on the single cell RNA sequencing data we identified 33 different cell types or states in the upper respiratory tract of these individuals including 21 immune and 12 epithelial cell subtypes (Fig. 1b, Extended Data Fig.1 a, b). We observed striking differences between the pediatric and adult study participants regarding the composition of the immune cell and epithelial cell compartment in the nasal mucosa. While immune cells were rarely detected in nasal samples from healthy adults, samples from SARS-CoV-2 negative children contained high amounts of almost each immune cell subset with an overall dominance of neutrophils (Fig. 1b, c, Extended Data Fig.2a). In adults, SARS-CoV-2 infection was associated with immune cell influx, while the proportion of immune and epithelial cells remained nearly stable in children (Fig. 1c, Extended Data Fig. 2a). Upon infection, children’s neutrophils showed an activated phenotype that was more pronounced than in infected adults, characterized by the enhanced expression of e.g. *CCL3* and *CXCR1/2* (Suppl. Table 4).

**Figure 1:**
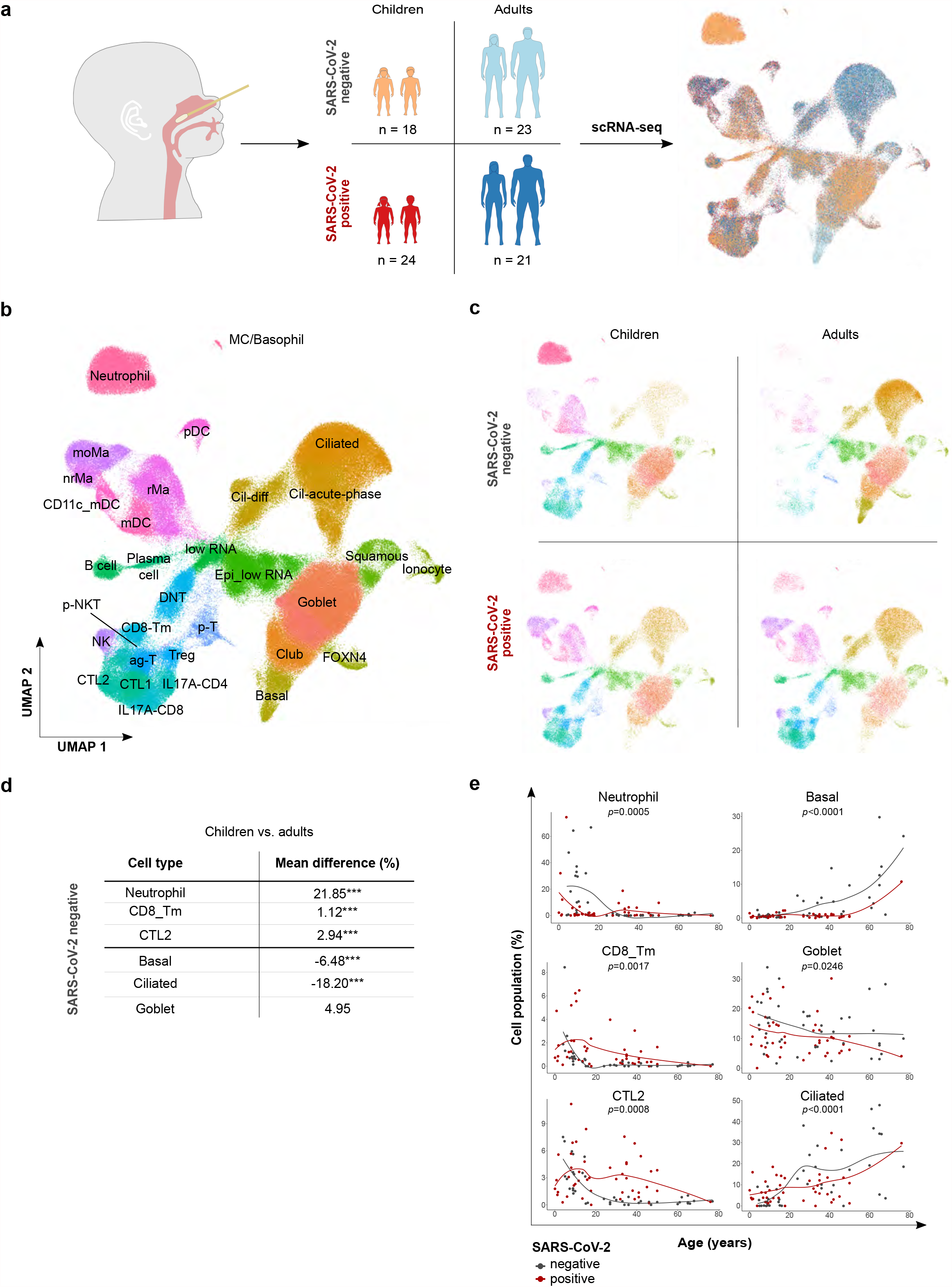
Age-dependent changes in cell composition of the upper airways. **(a)** Nasal samples of children (n=42) and adults (n=44) were collected from SARS-CoV-2 negative and SARS-CoV-2 positive (asymptomatic/mild/moderate COVID-19) individuals and subjected to scRNA sequencing. **(b)** UMAP showing all identified individual immune and epithelial cell types and states. **(c)** Scaled UMAPs displaying 45,000 cells per group reveal pronounced differences in the nasal cell composition of SARS-CoV-2 negative and positive children and adults **(d)** Table showing all cell types/states significantly different between children and adults in non-infected individuals. Given are mean differences in percent, positive values indicate a higher number of cells of the respective cell population in children compared to adults. Comparisons by Kruskal-Wallis test followed by Dunn’s two-sided post hoc comparison (*** *P* < 0.0001), also see Extended Data Figure 2a. **(e)** Scatter plots representing changes of specific immune and epithelial cells over time. Depicted are the percentages of the respective cell type/state with respect to all cells for immune cells (left) or epithelial cells (right) of each individual. SARS-CoV-2 negative individuals are indicated in grey, SARS-CoV-2 positive individuals in red. Lines represent curve fitting results by local polynomial regression (LOESS). P-value from linear regression analysis (Benjamini–Hochberg adjusted two-tailed).

Interestingly, many of the epithelial cell populations showed a clear age dependency with, e.g., goblet cells decreasing and ciliated cells increasing with age (Fig. 1d, e). A recent complementary study analyzed the cell composition of the nasal mucosa in healthy and SARS-CoV-2 infected children based on bulk RNA-Seq and cell deconvolution methods. They were unable to identify children-specific goblet cells, but rather described that samples from healthy children were dominated by a ciliated cell signature highlighting the limitations of bulk RNA approaches ^10^.

The expression of the SARS-CoV-2 entry receptor *ACE2* and the entry-associated proteases *TMPRSS2, FURIN, CTSB, CTSL*, and *CTSV* was similar between children and adults and not up-regulated by mild/moderate COVID-19 compared to the uninfected status (Extended Data Fig.2b). Hence, these viral entry factors cannot explain the differences in SARS-CoV-2 pathophysiology between children and adults.

SARS-CoV-2 is a positive-strand RNA virus with a very high rate of replication ^11, 12^. Hence, the control of SARS-CoV-2 infection requires an optimal early and coordinated innate antiviral immunity. This response is activated by various PRRs. Recently, mounting evidence has been generated in support of MDA5 (*IFIH1*) as the major PRR for SARS-CoV-2 in epithelial cells with RIG-I (*DDX58*) possibly playing an additional, but minor role ^13, 14^ (own unpublished data). An important enhancer of viral RNA sensing by MDA5 is LGP2 (*DHX58*) ^15^. Importantly, PRRs, in particular MDA5 and LGP2, are only weakly expressed in many epithelial cell types but are profoundly upregulated by positive feedback regulation upon viral infection of the cell or by paracrine exposure to type I or III interferon (IFN). The dynamics of this feedback regulation are crucial for the successful control of an infecting virus (Fig. 2a). The importance of the PRR/IFN axis for the successful resolution of SARS-CoV-2 infection was recently demonstrated by clinical studies finding a strong association between genetic polymorphisms at various loci of the PRR/IFN system with an increased risk of severe COVID-19 ^16^. Similarly, and affecting even a much broader fraction of patients, autoantibodies directed against type I IFNs have been shown to occur at a remarkably high frequency in patients suffering from severe COVID-19 ^17^.

**Figure 2:**
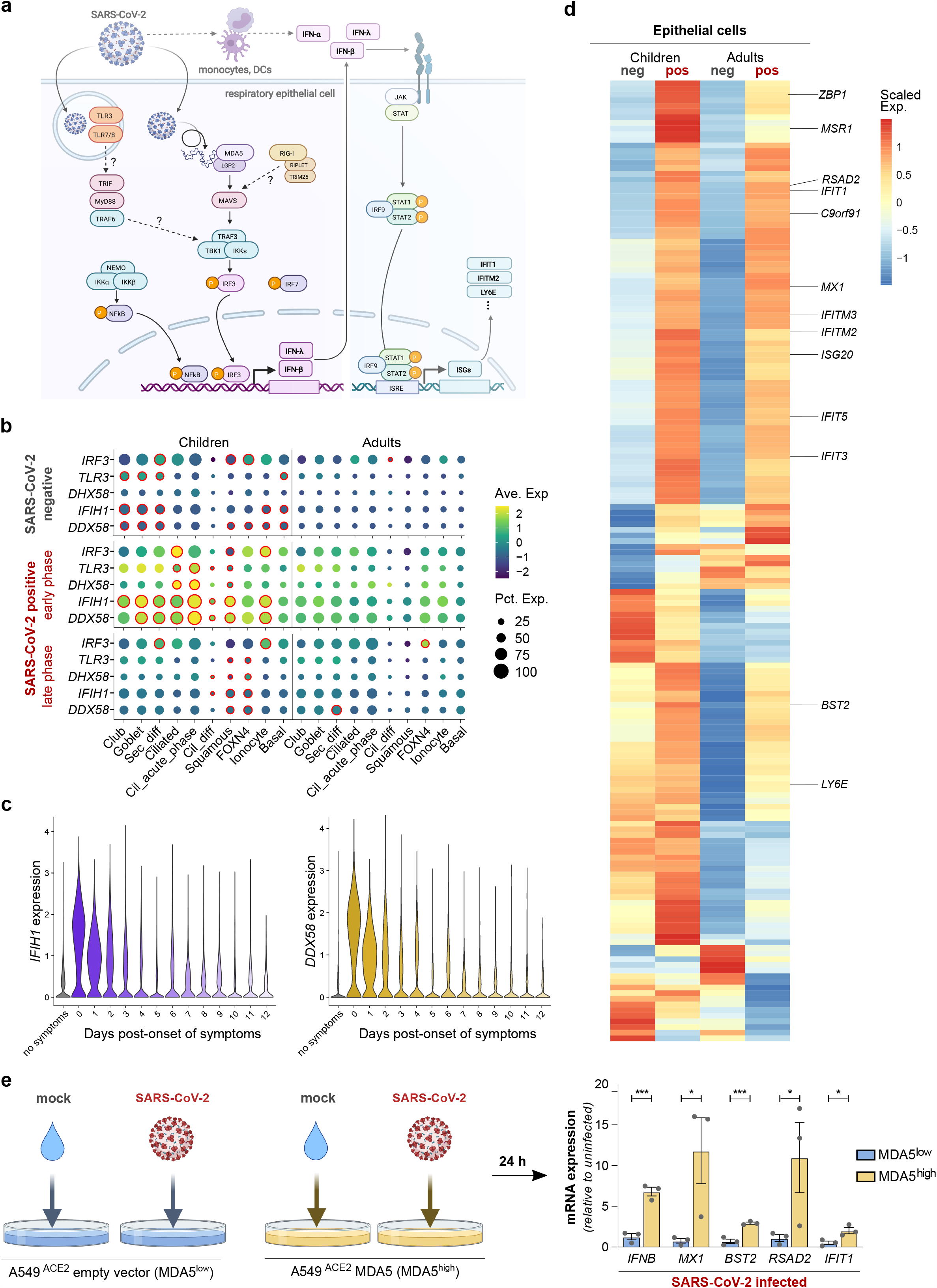
Enhanced viral sensing in children’s epithelial cells. **(a)** Schematic representation of epithelial response to SARS-CoV-2 infection including genes involved in virus sensing and subsequent interferon response. **(b)** Dot plots depicting the expression of viral sensing genes in epithelial cells of children and adults. Each significant increase comparing SARS-CoV-2 negative children (n=18) with SARS-CoV-2 negative adults (n=23), and SARS-CoV-2 positive children during the early (dps≤4, n=11) or late infection phase (dps 5-12, n=11) with SARS-CoV-2 positive adults amid early (n=13), or late (n=8) infection phase, respectively, is marked by a red circle (Benjamini–Hochberg adjusted two-tailed, Wilcoxon *P* < 0.05). Ave. Exp., average gene expression; Pct. Exp., percentage of cells expressing the gene. **(c)** Violin plots showing expression of prototypical virus sensing genes in epithelial cells of SARS-CoV-2 positive patients (n=45) in relation to days after first symptoms. **(d)** Heat map showing scaled expression of 171 representative ISGs in all epithelial cells during the early SARS-CoV-2 infection phase (dps ≤ 4). Only selected genes are annotated, for a completely annotated heat map see Extended Data FIG.3. **(e)** Differential expression of selected ISGs in an *in vitro* model comparing the response to SARS-CoV-2 infection in MDA5 (encoded by *IFIH1*) overexpressing A549^ACE2^ cells to empty vector controls. Bars show mean +/- SEM, *P<0.05, ***P<0.001 from one-tailed Student-t-test.

Notably, we found a significantly higher basal expression of the genes coding for RIG-I, MDA5 and LGP2 in epithelial cells in the upper respiratory tract of healthy children as compared to adults (Fig. 2b). This result suggests an increased ability of the respiratory mucosa of children to respond to viral infections, which is further supported by the highly increased amounts of innate immune cells in their upper airways (Fig. 1, Extended Data Fig. 2a). In epithelial cells of SARS-CoV-2 positive children and adults we observed a high expression of those genes (Figure 2c) in particular at the onset of COVID-19 symptoms that tended to decline until day 4 (day 0 – 4, referred to as early phase) and sustained at lower levels in the later disease phase (days 5 to 12 post symptom onset, referred to as late phase). It can be assumed that higher basal expression of these PRRs would permit immediate sensing of SARS-CoV-2 by MDA5/LGP2 in infected epithelial cells (Fig. 2a). Strikingly, children’s airway epithelial cells displayed increased expression of these PRR genes compared to the expression level of these genes in epithelial cells in SARS-CoV-2 positive adults (Fig. 2b), in particular in the early disease phase after symptom onset. From day 5 onwards, virus sensing is largely comparable between children and adults (Fig.2b, lower plot).

Following virus sensing, signaling through IRF3/NFkB leads to the expression of primary antiviral effectors, as well as antiviral cytokines such as IFN-β and IFN-λ (Fig. 2a). IFNs act on epithelial cells in an auto- and paracrine manner, further increasing MDA5/LGP2 responsiveness in the tissue and inducing a broad range of interferon-stimulated genes (ISG). While we were not able to detect the expression of type I and type III interferons themselves, ISGs showed an impressive activation pattern in epithelial cells of SARS-CoV-2 positive children, including many genes previously shown to exhibit strong antiviral activity against SARS-CoV-2, such as *LY6E* ^*18*^, *IFITM2*, and *BST2* ^*19*^. In all epithelial cells, and in particular in ciliated cells, the magnitude of ISG expression considerably surpassed that of infected adults in both the early and late infection phase (Fig. 2d, Extended Data Fig. 3) with a generally decreasing trend in the late phase (Extended Data Fig. 3).

To demonstrate a direct association between MDA5 expression levels and activation of ISGs upon SARS-CoV-2 infection, we established an *in vitro* model using the human lung epithelial cell line A549, which exhibits very low basal expression levels of MDA5 similar to expression levels found in nasal epithelial cells of healthy adults. As expected, based on inefficient MDA5 sensing in concert with rapid replication and expression of virus-encoded antagonists ^20^, only minute amounts of IFN and ISG transcripts were induced upon SARS-CoV-2 infection in these cells. However, in cells with moderately increased basal expression of MDA5 by lentiviral transduction, permitting efficient virus sensing prior to the expression of antagonists, we observed a significant induction of the expression of *IFNB* and key ISGs including *MX1, BST2* (Tetherin), *RSAD2* (Viperin) *and IFIT1* (Fig. 2e). These findings corroborate the central role of MDA5 expression prior to infection for sensing SARS-CoV-2 and inducing a swift and robust ISG response.

Studying immune-epithelial cell interactions revealed a stronger immune-epithelial cell cross-talk in children vs. adults particularly before infection. Among immune cells non-resident (nrMa) and monocyte-derived macrophages (moMa) as well as CD11c^+^ dendritic cells (CD11c_mDCs) were most interactive (Fig. 3a). These immune cell subsets showed a higher activation status in children as demonstrated by an increased expression level of several cytokine and chemokine-coding genes such as *IL1B, IL8, TNF, CCL3 and CCL4* (Fig. 3b). We furthermore observed an enhanced expression of *IFIH1* in moMas, nrMas, and CD11c_mDCs in SARS-CoV-2 infected children and a significant increase of *TLR2* in moMa, nrMa in the early phase of infection, suggesting that these cells might play an additional role in virus sensing and IFN production. This is further underlined by the fact that moMa, nrMa and CD11c_mDCs of SARS-CoV-2 negative children expressed *IFIH1* and *TLR2* at higher levels than adults.

**Figure 3:**
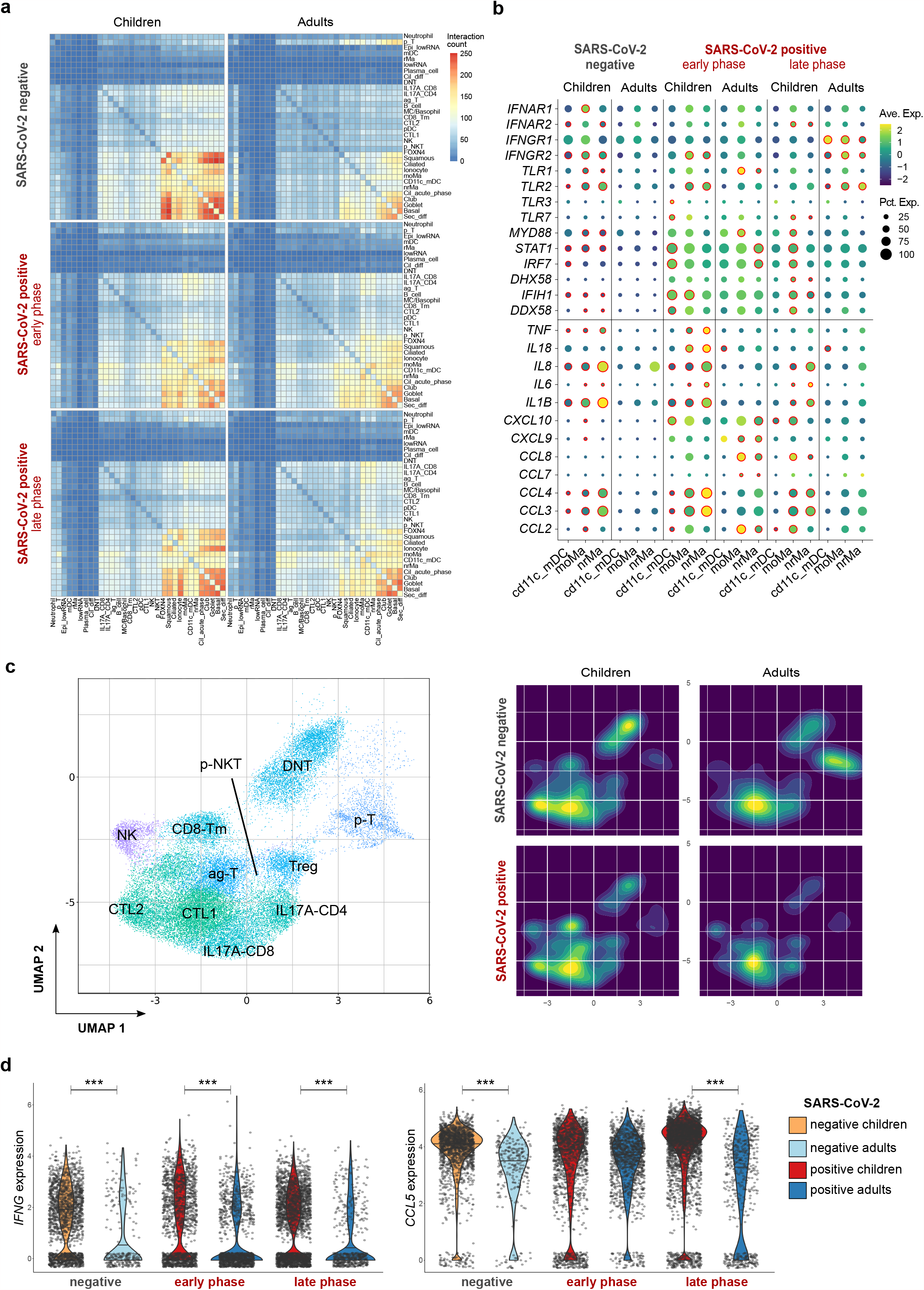
Differential immune response in children and adults. **(a)** Heat maps depicting cell–cell communications between all identified cell types derived from linear-scaled ligand– receptor interaction counts in children (left, n = 18 SARS-CoV-2 negative, n=11 SARS-CoV-2 positive early infection phase (dps ≤ 4), n=11 SARS-CoV-2 positive late infection phase (dps 5-12)) and adults (right, *n* = 23 SARS-CoV-2 negative, n=13 SARS-CoV-2 positive early infection phase, n=8 SARS-CoV-2 positive late infection phase) **(b)** Dot plots depicting expression of genes involved in anti-viral and cytotoxic response of myeloid dendritic cells and different macrophage populations. Each significant increase comparing SARS-CoV-2 negative children (n=18) with SARS-CoV-2 negative adults (n=23) and SARS-CoV-2 positive children during the early (n=11) or late (n=11) infection phase with SARS-CoV-2 positive adults amid early (n=13), or late (n=8) infection phase, respectively is marked by a red circle (Benjamini– Hochberg adjusted two-tailed Wilcoxon, *P* < 0.05). Ave. Exp., average gene expression; Pct. Exp., percentage of cells expressing the gene. **(c)** UMAP displaying the different T/NK cell subpopulations in the nose of children and adults. In comparison, scaled density plots are shown indicating the proportion of the different T/NK cells in children and adults separated by their SARS-CoV-2 infection status. Yellow represents high density, blue represents low density. **(d)** Violin plots showing expression of representative immune mediators of the CTL2 subpopulation comparing SARS-CoV-2 negative children (n=18) with SARS-CoV-2 negative adults (n=23) and SARS-CoV-2 positive children during the early (n=11) or late (n=11) infection phase with SARS-CoV-2 positive adults amid early (n=13), or late (n=8) infection phase, respectively. Each dot represents one cell. Plots show median, \*\*\**P* < 0.001 derived from two-tailed Wilcoxon comparison.

Apart from the up-regulated cell-intrinsic antiviral capacity of airway epithelial cells, macrophages and DCs, we found specific patterns of immune cell subpopulations in children vs. adults. Among others we identified a subpopulation of *KLRC1* (NKG2A)^+^ cytotoxic T cells (CTL2) occurring predominantly in children (Fig. 3c). NKG2A is a lectin-like inhibitory receptor on cytotoxic T cells playing a role in limiting excessive activation, preventing apoptosis and sustaining the virus-specific CD8+ T cell response ^21^. Already without viral infection, this CD8 cytotoxic T cell subset was characterized by a strong expression of cytotoxic mediators (Fig. 3d, Extended Data Fig. 4a, Suppl. Table 3). Furthermore *IFNG* was highly expressed in these cells when comparing SARS-CoV-2 negative children to adults. Upon infection, children were characterized by a significantly higher expression of *IFNG* compared to adults both in the early phase and in the later phase of infection. Similarly, the potent chemoattractant *CCL5* was increased in children compared to adults with or without infection (Fig. 3d). The cytotoxic potential and the predominance of this cytotoxic T cell subset necessary for efficient killing of virus-infected cells provides further evidence for a better anti-virus response in children compared to adults. In addition, SARS-CoV-2 infected children showed a distinct CD8^+^ T cell population (CD8-Tm) with a memory phenotype that was almost absent in adults (Figure 3c, EDF4). It remains unclear whether these cells are beneficial for protection of the children against future reinfection.

Taken together our data provide clear evidence that the epithelial and immune cells of the upper airways (nose) of children are pre-activated and primed for virus-sensing. This is likely a general feature of the children’s mucosal immune response, but of particular relevance for SARS-CoV-2. Very recently, scRNASeq of Chikungunya virus-infected fibroblasts showed an extremely narrow window of opportunity for the cells to express IFNs before viral protein production shuts the antiviral system off ^22^. This likely also explains the differences between SARS-CoV-2 and other respiratory viruses including RSV, influenza A virus (IAV), or SARS-CoV-1 in terms of the induced host response. SARS-CoV-2 is characterized by extensive intracellular replication and a remarkable absence of IFN-production and -secretion. On the other hand, SARS-CoV-2 is highly sensitive to treatment with IFNs prior to or after infection as shown in lung epithelial cells, even more so than SARS-CoV-1 ^20, 23^. Primed virus sensing and a pre-activated innate immune response in children leads to efficient early production of IFNs in the infected airways, likely mediating substantial antiviral effects mirroring those observed *in vitro* in IFN-(pre)treated cells. Ultimately, this may lead to reduced virus replication and faster clearance in children. In fact, several studies already showed that children are much quicker in eliminating SARS-CoV-2 compared to adults consistent with the concept that they shut down viral replication earlier ^24-27^. For other respiratory viruses, such as RSV and IAV, that more efficiently induce an IFN response by themselves, a pre-activated innate immune response may be less relevant. The enhanced innate antiviral capacity in children together with the high IFN sensitivity of SARS-CoV-2 may explain why children are better able to control early-stage infection as compared to adults and therefore have a lower risk of developing severe COVID-19.

## Data Availability

Due to potential risk of de-identification of pseudonymized RNA sequencing data the raw data will be available under controlled access in the EGA repository, [will be added upon completion of peer review/ acceptance]. Count and metadata tables (patient-ID, sex, age, cell type, QC metrics per cell) can be found at FigShare: [will be added upon completion of peer review/ acceptance]. In addition, these data can be further visualized and analyzed in the Magellan COVID-19 data explorer at https://digital.bihealth.org [will be publicly available upon completion of peer review/ acceptance].

## Methods

### Patient Recruitment and Ethics Approval

Individuals of three different cohorts were included in this study. Patients of the prospective observational cohort study Pa-COVID-19 ^28^ and its study arm RECAST (Understanding the increased **RE**silience of **C**hildren compared to **A**dults in **S**ARSCoV-2 infec**T**ion) were enrolled between August 2020 and June 2021 at Charité – Universitätsmedizin Berlin. Further patients were recruited in the prospective SC2-Study ^8, 9^ at University Hospital Leipzig between March 2020 and May 2021. Written informed consent was given by all patients and/or their parents prior to inclusion. All three studies were conducted in accordance with the Declaration of Helsinki and approved by the respective Institutional Review Boards (Pa-COVID-19/ RECAST: EA2/066/20, SC2: 123/20-ek).

#### Patient cohort

From the three cohorts patients classified as asymptomatic, mild, or moderate based on WHO guidelines ^29^ of COVID-19 severity were enrolled. In total, we analyzed nasal swabs of 45 confirmed SARS-CoV-2 positive patients comprising 24 children with ages ranging from 4 weeks to 17 years (median age 9.0 ± 5.6 years, 10 females, 14 males) and from 21 adults between 21 and 76 years (median age 39.0 ± 10.4 years, 12 females, 9 males). None of the children were hospitalized, but all were in domestic quarantine. Additionally, nasal swabs of 42 healthy, SARS-CoV-2 negative controls from 18 children between 4 and 16 years (median age 9.0 ± 3.8 years, 8 females, 10 males) and from 23 adults between 24 and 77 years were included (median age 46.0 ± 16.3 years, 13 females, 10 male, Suppl. Table 1 & 2). All negative controls were examined for possible exposure to SARS-CoV-2 by detailed anamnesis. In the cases of known SARS-CoV-2 exposures additional serological testing was conducted at the time of sampling and after 14 days or follow-up interviews were conducted to ensure that the participant as well as their household members showed no signs of infection for at least two consecutive weeks and that any routine testing yielded negative results.

#### Real-Time Reverse Transcription PCR for SARS-CoV-2

RNA was extracted by using the MagNA pure 96 DNA and viral NA small volume Kit (Roche, https://www.roche.com) on a MagNA Pure 96 System as recommended by the manufacturer. Real time reverse transcription PCR (rRT-PCR) was performed targeting the envelope (E) gene and nucleocapsid (N) gene on the Roche Light Cycler 480 system (Tib-Molbiol, https://www.tib-molbiol.de).

### Obtaining a single cell suspension from human nasal swabs, preparation for single cell RNA sequencing and subsequent pre-processing of the raw data

Sample processing, single-cell and library preparation, and data analysis were performed as documented previously ^8, 9^. Briefly, fresh nasopharyngeal swabs were transferred into cold DMEM/F12 medium (Gibco, 11039) and within one hour processed further. Under biosafety S2 an equal volume of 13 mM DTT (AppliChem, A2948) was added to each sample. To achieve higher cell numbers, the solution was slowly pipetted up and down and the swab was dipped roughly 20 times into the medium. Following incubation at 37°C, 500 rpm for 10 minutes on a thermomixer, samples were centrifuged at 350xG at 4°C for 5 minutes and the supernatant slowly removed. If the pellet showed any sign of red blood cells (RBC), it was resuspended in 1x PBS (Sigma-Aldrich, D8537), treated with RBC Lysis Buffer (Roche, 11814389001) at 25°C for 10 minutes and centrifuged at 350xG at 4°C for 5 minutes. If samples were not processed immediately, the cell pellet was resuspended in DMEM/F12 supplemented with 20% FBS (Gibco, 10500) and 10% DMSO (Sigma-Aldrich, D8418) and frozen at -80°C. For the library preparations cells were thawed at 37°C, centrifuged at 350xG at 4°C for 5 minutes and further processed according to the protocol. To obtain a single cell suspension Accutase (Thermo Fisher, 00-4555-56) was added to the pellet and the solution incubated at room temperature for 10 minutes with carefully pipetting the cells after 5 minutes. The incubation was stopped by adding DMEM/F12 supplemented with 10% FBS and centrifugation at 350xG at 4°C for 5 minutes. Subsequently, the supernatant was removed and the cell pellet was resuspended in 1x PBS (volume was adjusted to the size of the cell pellet). The suspension was cleared of any cell debris using a 35 μm cell strainer (Falcon, 352235) and subsequently, cells were counted with a disposable Neubauer chamber (NanoEnTek, DHC-N01). Cell suspension was diluted to allow loading of 17,500 cells per sample. A single cell and unique barcode emulsion was achieved by mixing the diluted cells with the master mix and loading them on the chip together with the Gel Beads and Partitioning Oil using the 10x Genomics Single Cell 3’
s GEM, Library and Gel Bead Kit v3.1 (10x Genomics; PN 1000120; PN 1000121; PN 1000213) and loading the chip into the 10x Chromium Controller. The following reverse transcription, clean-up and cDNA amplification, as well as the library preparation were performed according to the manufacturer’s manual. Notably, to make sure that the virus was inactivated we prolonged the incubation at 85°C during the reverse transcription to 10 minutes. Final 3’ RNA libraries were pooled for sequencing either on a S2 or S4 flow cell (S2: up to 13 samples, S4: up to 24 samples) and sequenced on the NovaSeq 6000 Sequencing System (Illumina, paired-end, single-indexing).

Single-cell datasets were aligned and preprocessed using cellranger 3.0.1. A custom human hg19 reference genome (10x Genomics, version 3.1.0) with the SARS-CoV-2 genome (Refseq-ID: NC_045512) added as additional chromosome was used. For downstream analysis Seurat 3.2.2 was used. Cells with less than 3 genes and cells with more than or equal to 15% mitochondrial reads or less than 200 genes expressed were discarded. To remove doublets a cutoff for the number of UMIs and genes was determined manually per sample.

Samples were merged and exported as csv files containing counts and metadata and imported into scanpy 1.6.0 (Wolf et al., 2018). Following normalization to 10,000 reads per cell, expression values were log transformed and highly variable genes were calculated, which were used as basis for the following preprocessing steps. The data were scaled, PCA transformed and aligned using harmony 0.0.5 ^30^ based on 100 principal components. Further alignment was performed using bbknn ^31^ based on 50 prealigned principal components, 10 trees, 3 neighbors within batch and a trim setting of 85. Based on the integrated data UMAP embedding and leiden ^32^ clustering were performed. This clustering was used as a basis for subclustering of immune and epithelial cell populations with the same algorithm. Clusters of epithelial ^33-35^ and immune ^36-38^cell populations were assigned to cell types/ stages according to the expression level of different marker genes (Extended Data Fig. 1, Extended Data Fig. 4). The T cell and macrophage/DC clusters were sub-clustered and then further refined manually.

The object was stored as h5ad, converted to h5seurat using SeuratDisk version 0.0.0.9014 and imported back into R.

In total, 268,745 cells were included in the data set. Cell numbers between the groups were equally distributed (neg. children 51,595; neg. adults 62,701; pos. adults 51,500) with the exception of the group of positive children containing higher cell numbers (102,949). Different samples contributed varying numbers of cells. The percentage of contribution of each sample to its study group were compared using a Kruskal-Wallis test, which did not indicate significant differences between groups (p=0.2).

To enable visual comparisons between UMAPs of different groups, equal numbers of cell (45,000) per group were randomly sampled using SubsetData function in Seurat.

Putative cell-cell interactions were quantified using CellPhoneDB version 2.1.2 using default settings ^39^. In order to reduce the influence of individual samples contributing a larger number of cells and to speed up computation, we capped the number of cells per sample at randomly sampled 2,000 cells. This was done using the SubsetData function in Seurat.

### Identification of ISG gene set

For the analysis of PRR/IFN responses, a gene set of the most prominent ISGs expressed by lung epithelial cells was assembled. As described previously ^9^, we treated A549 epithelial cells with a mix of IFN-β and IFN-λ for 2, 8 or 24 h, and analyzed transcript levels by microarray analyses using the Illumina Human HT-12 Expression Beadchip platform at the genomic and proteomics core facility at DKFZ. We identified ISGs as exhibiting a log-2-fold-change > 0.8 at any time point, yielding 183 genes. We further included ISGs described to exhibit strong anti-SARS-CoV-2 activity (65 top-scoring genes) if not already included in our list. This eventually yielded a gene set of 217 genes also expressed in our scRNA-Seq.

### SARS-CoV-2 infection of MDA5-expressing A549 cells

A549 cells stably transduced with a lentiviral vector expressing human *IFIH1* under the control of the murine ROSA26 promoter (termed A549 MDA5^high^ in Fig. 2e) were kindly provided by Nadine Gillich and Ralf Bartenschlager. We transduced A549 (termed MDA5^low^) and A549 MDA5^high^ cells using lentiviral vectors encoding human ACE2 and TMPRSS2 in order to make them permissive for SARS-CoV-2 infection; in order to ensure consistent ACE2/TMPRSS2 expression across experiments, transduction was freshly done 24 h prior to infection of cells. Infection with SARS-CoV-2 (strain BetaCoV/Germany/BavPat1/2020) was performed in our BSL3 facility at a MOI of 0.1, and cells were harvested at 24 h post infection. RNA was extracted using the Monarch Total RNA Miniprep Kit (New England Biolabs) and reverse transcribed by the High Capacity cDNA Reverse Transcription Kit (ThermoFisher Scientific). IFN-β and ISG transcript levels were then assessed by RT-PCR using the iTaq Universal SYBR Green Supermix (BioRad) on a BioRad CFX96 Real-Time PCR Detection System. Data is presented as mean +/- SEM of three biologically independent replicates.

### Statistics

Differential gene expression was calculated using “rank_genes_groups()” in scanpy version 1.6.0 and corrected for false discovery rates with statsmodels version 0.9.0 ^40^. Differences in cell type / stage compositions were assessed using the Kruskal-Wallis rank sum test followed by two-tailed Dunn’
ss post-hoc test. Age dependencies were calculate fitting a linear regression model corrected for the COVID-19 status and adjusting the F-test p-values using the Benjamini Hochberg method ^41^.

P-values for dot plots and violin plots were calculated using the Seurat function “FindMarkers()” based on the Wilcoxon test and corrected with the Benjamini Hochberg method^41^.

To test whether elevated MDA5 levels in A549 cells significantly increased IFNB and ISG induction upon infection, an unpaired one-tailed Student-*t-*test of three biologically independent repetitions was performed (GraphPad Prism v9.1).

## Data availability

Due to potential risk of de-identification of pseudonymized RNA sequencing data the raw data will be available under controlled access in the EGA repository, [will be added upon manuscript acceptance]. Count and metadata tables (patient-ID, sex, age, cell type, QC metrics per cell) can be found at FigShare: [will be added upon manuscript acceptance]. In addition, these data can be further visualized and analyzed in the Magellan COVID-19 data explorer at https://digital.bihealth.org [will be publicly available upon manuscript acceptance].

## Code availability

No custom code was generated/used during the current study.

## Acknowledgement

We thank all patients of the RECAST, Pa-COVID-19, and SC2 studies for kindly donating nasal samples and clinical data. This study was supported by the BIH COVID-19 research program and the fightCOVID@DKFZ initiative, the European commission (ESPACE, 874719, Horizon 2020), the German Federal Ministry for Education and Research (BMBF) - funded de.NBI Cloud within the German Network for Bioinformatics Infrastructure (de.NBI; 031A537B, 031A533A, 031A538A, 031A533B, 031A535A, 031A537C, 031A534A, 031A532B), the BMBF-funded Medical Informatics Initiative (HiGHmed, 01ZZ1802A - 01ZZ1802Z), the BMBF-funded projects 01IK20337, 82DZL0098B1, the German Research Foundation (Deutsche Forschungsgemeinschaft, DFG)-funded CRC-TR 84 B08, CRC-1449 Z01, CRC-TR 179 TP11 and the DFG COVID-19 focus funding project BI1693/2-1.

## Authors contributions

ST, SL, IL, MAM and RE conceived, designed, and supervised the project.

BS, MR, JR, LS, VMC, and MAM designed the RECAST cohort

JR, SS, LS, SvL recruited the patients, provided the human specimens, clinical data and annotation of the patients.

JeLo, AS, JoLi, MaMe, RLCh performed scRNA experiments.

VGM and MB performed and analyzed the *in vitro* experiments.

JeLo, LT, SL, RLCh analyzed data.

BS, LS, SvL, MVC, MR, CC, MB and MAM contributed with discussion of the results.

BT and SK provided technical and experimental support.

JeLo, SL, ST, MB, RE, and IL wrote the manuscript, all authors read, revised, and approved the manuscript.

## Extended Figure Legends

*Extended Data Figure 1:* **Cell type marker genes. (a-b)** Dot plots depicting average and percent expression of genes used to classify **(a)** immune and **(b)** epithelial cells. Pct. Exp = percentage of cells expressing the gene, Ave. Exp. = average gene expression.

*Extended Data Figure 2:* **Cell composition in children and adults and ACE2 expression. (a)** Distribution of all cell types/ states in children and adults separated by SARS-CoV-2 infection status. Given are percentages of the total number of cells. Comparisons by Kruskal-Wallis test followed by Dunn’s two-sided post hoc comparison (*significance of comparison between children and adults, P < 0.05). **(b)** Violin plots show distribution of the SARS-CoV-2 entry receptor *ACE2* expression and the gene expression of associated proteases in epithelial cells of children and adults either with (n=24 children, n=21 adults) or without SARS-CoV-2 infection (n=18 children, n=23 adults). Plots show median. No significant differences were observed.

*Extended Data Figure 3:* **Epithelial IFN-response**. Heat maps depicting scaled expression (zero mean, unit variance) of known IFN-response genes in all epithelial cells and separately in ciliated cells across conditions during early and late SARS-CoV-2 infection phase (early: dps ≤ 4: n=11 children, n=13 adults; late dps >4: n=11 children, n=8 adults). Genes were filtered according to a minimum fold change of 1.5 between the lowest and highest expressing group, with the gene being selected according to their differential expression in all epithelial cells. Genes not meeting the criterion were greyed out to avoid inflating minor differences as a consequence of the scaling performed. Scaled Exp. = scaled expression

*Extended Data Figure 4:* **Characteristics of NK and T-cells**. Expression profile of cytotoxic and aging-related genes in the NK and T-cell subtypes derived from n=86 nasal swap samples distinguishing the children’s and adults’ immune profile. Ave. Exp. = average gene expression, Pct. Exp. = percentage of cells expressing the gene.

