## Supplement for "Pre-activated anti-viral innate immunity in the upper airways controls early SARS-CoV-2 infection in children"

### EDF1

**a**

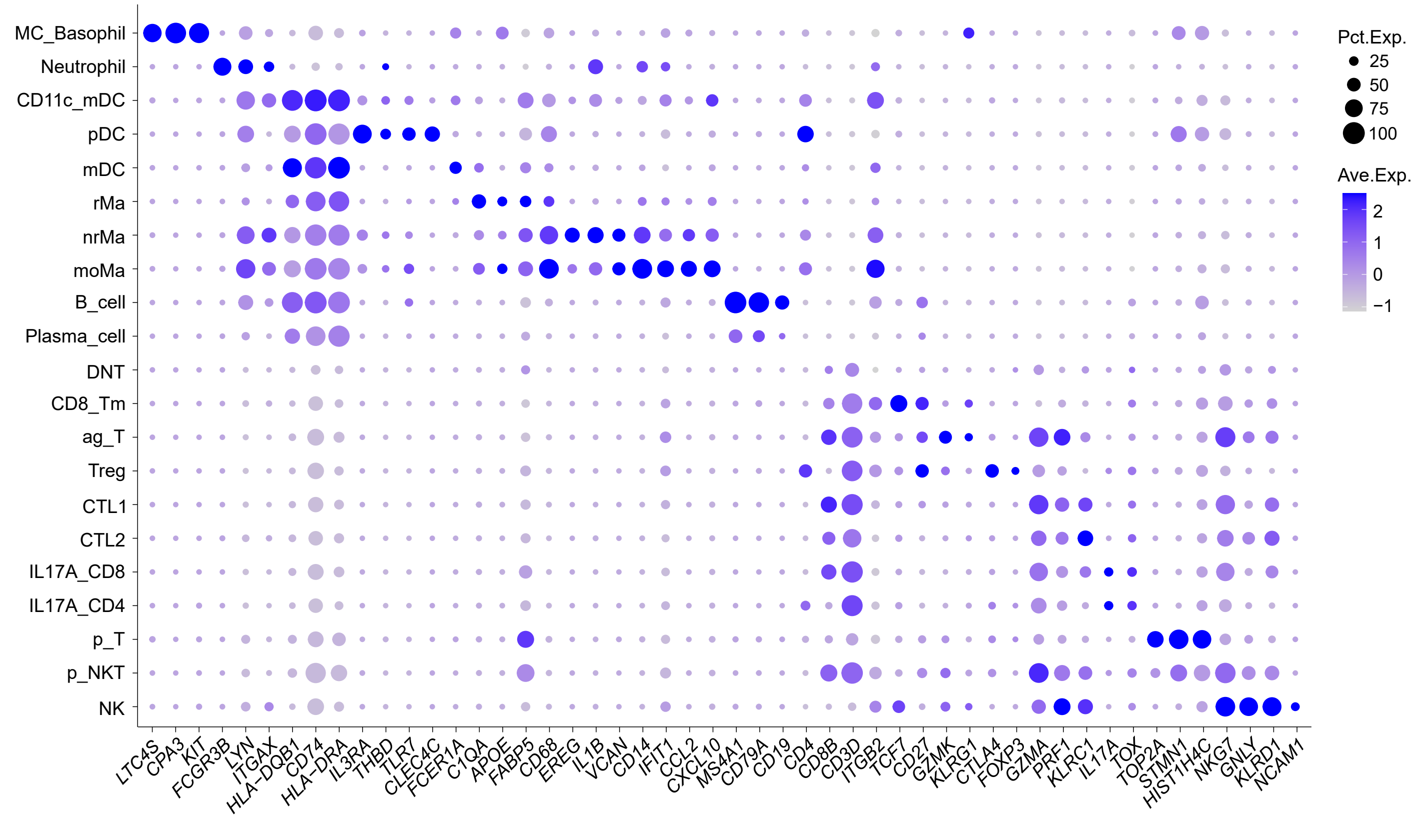
MC_Basophil
Neutrophil
CD11c_mDC
pDC
mDC
rMa
nrMa
moMa
B_cell
Plasma_cell
DNT
CD8_Tm
ag_T
Treg
CTL1
CTL2
IL17A_CD8
IL17A_CD4
p_T
p_NKT
NK
LTC4S CPA3 KIT FCGR3B LYN ITGAX HLA-DQB1 CD74 HLA-DRA IL3RA THBD TLR7 CLEC4C FCER1A C1QA APOE FABP5 CD68 EREG IL1B VCAN CD14 IFIT1 CCL2 CXCL10 MS4A1 CD79A CD19 CD4 CD8B CD3D ITGB2 TCF7 CD27 GZMK KLRG1 CTLA4 FOXP3 GZMA PRF1 KLRC1 IL17A TOX TOP2A STMN1 HIST1H4C NKG7 GNLY KLRD1 NCAM1
Pct.Exp.
25
50
75
100
Ave.Exp.
2
1
0
−1

**b**

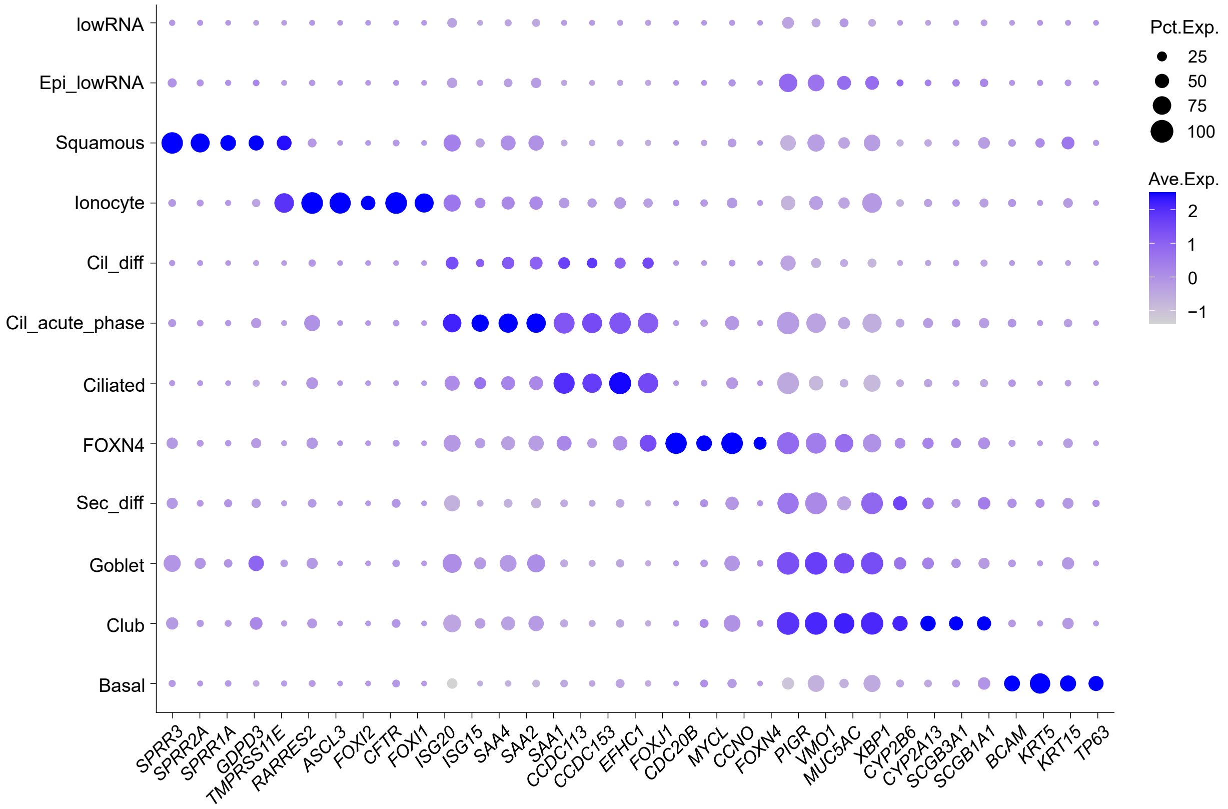
lowRNA
Epi_lowRNA
Squamous
Ionocyte
Cil_diff
Cil_acute_phase
Ciliated
FOXN4
Sec_diff
Goblet
Club
Basal
SPRR3 SPRR2A SPRR1A GDPD3 TMPRSS11E RARRES2 ASCL3 FOXI2 CFTR FOXI1 ISG20 ISG15 SAA4 SAA2 SAA1 CDC113 CCDC153 EFHC1 FOXJ1 CDC20B MYCL CCNO FOXN4 PIGR VMO1 MUC5AC XBP1 CYP2B6 CYP2A13 SCGB3A1 SCGB1A1 BCAM KRT5 KRT15 TP63
Pct.Exp.
25
50
75
100
Ave.Exp.
2
1
0
−1

### EDF2

## a

| | SARS-CoV-2 negative: Children | SARS-CoV-2 negative: Adults | SARS-CoV-2 positive: Children | SARS-CoV-2 positive: Adults |
|---|---|---|---|---|
| MC_Basophil | 0.25 | 0.18 | 0.15 | 0.57 |
| Neutrophil | 22.14*** | 0.28 | 6.04 | 3.01 |
| mDC | 2.66** | 0.85 | 1.99 | 1.72 |
| CD11c_mDC | 0.49 | 0.4 | 1.63 | 1.28 |
| pDC | 0.48 | 0.2 | 1.51 | 0.95 |
| rMa | 5.66** | 1.47 | 6.74 | 6.93 |
| nrMa | 0.41 | 0.23 | 2.14 | 1.76 |
| moMa | 0.78 | 0.4 | 5.22 | 4.88 |
| NK | 0.48** | 0.09 | 1.16 | 0.93 |
| p_NKT | 0.06* | 0.0 | 0.15 | 0.1 |
| p_T | 0.72 | 1.06 | 1.25 | 1.09 |
| ag_T | 1.01*** | 0.18 | 1.8 | 1.79 |
| Treg | 0.73** | 0.18 | 0.99 | 0.98 |
| DNT | 3.32*** | 0.56 | 2.79 | 2.13 |
| CD8_Tm | 1.21*** | 0.09 | 2.01 | 0.84 |
| IL17A_CD8 | 1.85*** | 0.36 | 1.89 | 1.66 |
| IL17A_CD4 | 1.08** | 0.4 | 1.27 | 1.47 |
| CTL2 | 3.31*** | 0.37 | 3.49 | 2.88 |
| CTL1 | 2.48** | 0.76 | 2.85 | 3.71 |
| Plasma_cell | 1.45*** | 0.1 | 0.8 | 0.42 |
| B_cell | 1.18** | 0.23 | 1.44 | 1.53 |
| immune_total | 51.74*** | 8.41 | 47.32 | 40.62 |
| lowRNA | 2.58 | 2.14 | 2.24 | 1.35 |
| Epi_lowRNA | 9.31 | 11.44 | 8.89 | 8.43 |
| Squamous | 2.18 | 6.39 | 1.8 | 4.93 |
| Ionocyte | 0.51 | 0.66 | 1.07 | 0.82 |
| FOXN4 | 0.66 | 0.63 | 0.81 | 0.51 |
| Basal | 0.68*** | 7.15 | 0.64 | 1.07 |
| Cil_diff | 1.74** | 4.71 | 4.08 | 5.11 |
| Cil_acute_phase | 1.87*** | 10.26 | 5.87 | 8.12 |
| Ciliated | 2.4*** | 20.59 | 6.79 | 11.85 |
| Sec_diff | 4.09** | 7.8 | 3.5 | 3.88 |
| Goblet | 16.89 | 11.94 | 12.71 | 9.59 |
| Club | 5.35 | 7.85 | 4.27 | 3.72 |

## b

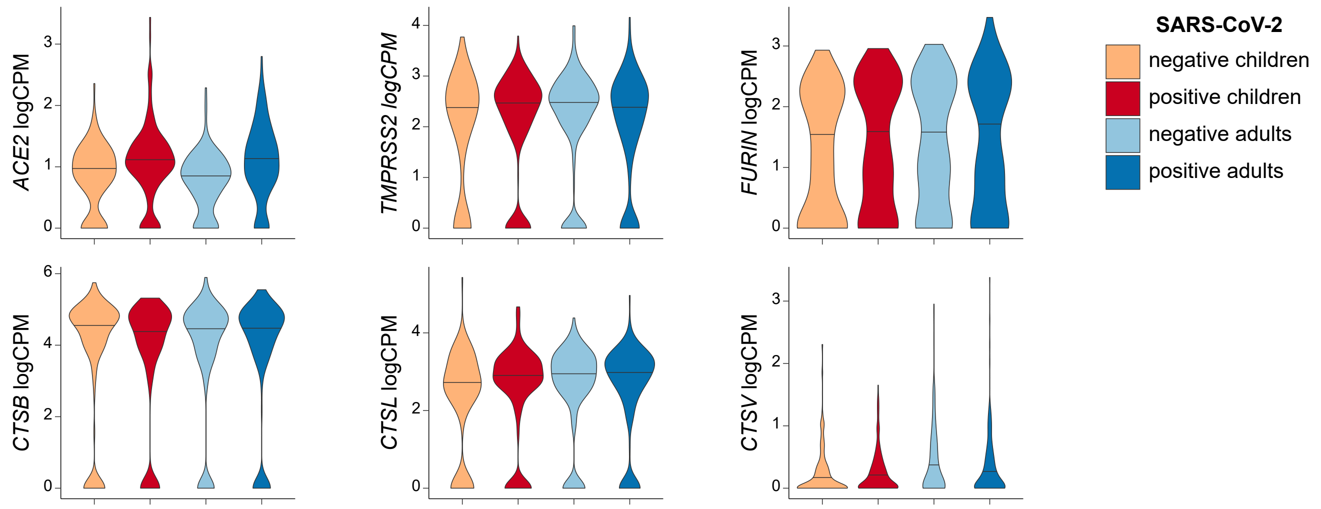

### EDF3

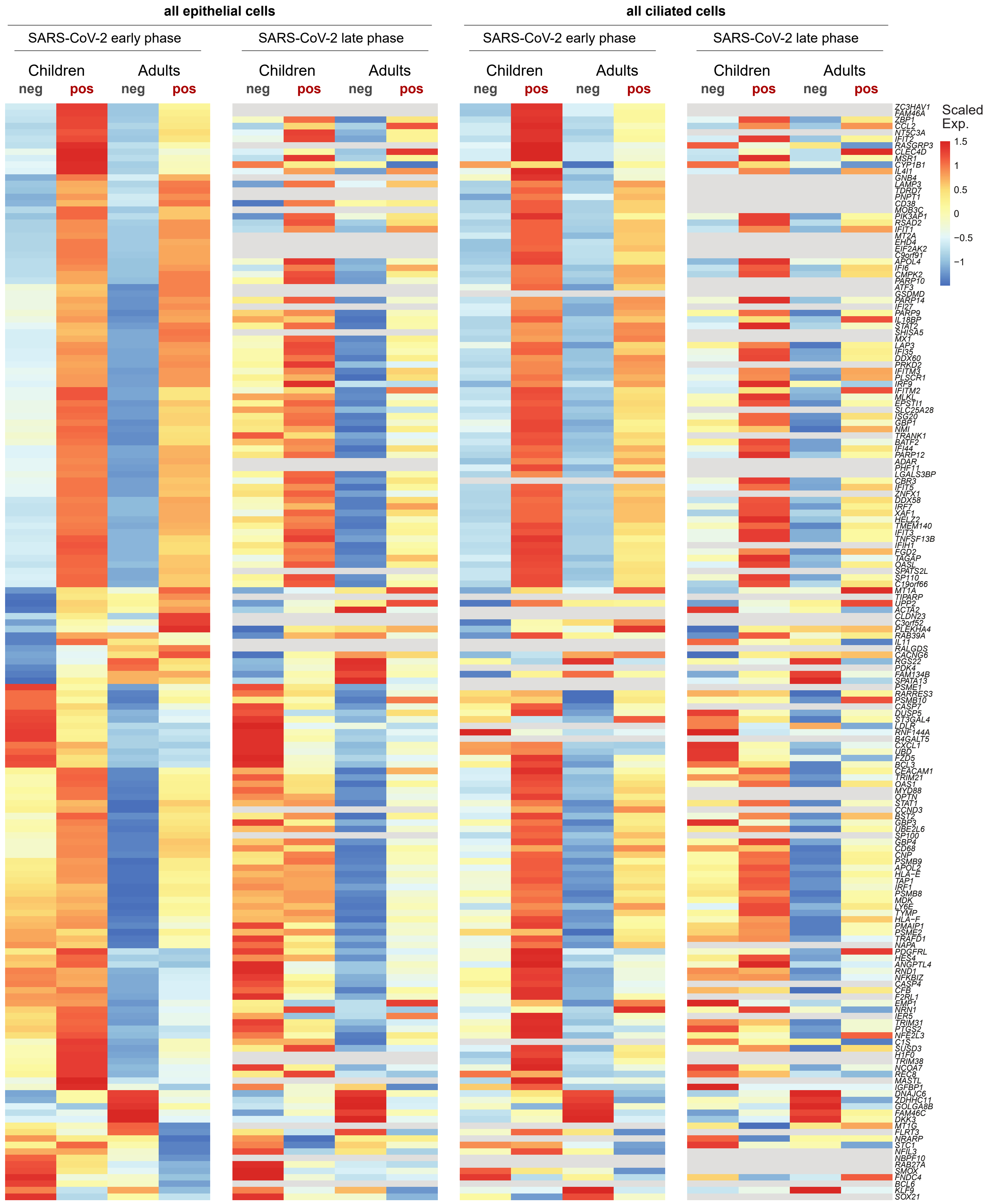
all epithelial cells
all ciliated cells
SARS-CoV-2 early phase
SARS-CoV-2 late phase
SARS-CoV-2 early phase
SARS-CoV-2 late phase
Children
Adults
Children
Adults
Children
Adults
Children
Adults
neg
pos
neg
pos
neg
pos
neg
pos
neg
pos
neg
pos
neg
pos
neg
pos
Scaled Exp.
1.5
1
0.5
0
−0.5
−1
ZC3HAV1
FAM46A
ZBP1
CCL2
NT5C3A
IFIT2
RASGRP3
CLEC4D
MSR1
CYP1B1
IL4I1
GNB4
LAMP3
TDRD7
PNPT1
CD38
MOB3C
PIK3AP1
RSAD2
IFIT1
MT2A
EHD4
EIF2AK2
C9orf91
APOL4
IFI6
CMPK2
PARP10
ATF3
GSDMD
PARP14
IFI27
PARP9
IL18BP
STAT2
SHISA5
MX1
LAP3
IFI35
DDX60
PRKD2
IFITM3
PLSCR1
IRF9
IFITM2
MLKL
EPSTI1
SLC25A28
ISG20
GBP1
NMI
TRANK1
BATF2
IFI44
PARP12
ADAR
PHF11
LGALS3BP
CBR3
IFIT5
ZNFX1
DDX58
IRF7
XAF1
HELZ2
TMEM140
IFIT3
TNFSF13B
IFIH1
FGD2
TAGAP
OASL
SPATS2L
SP110
C19orf66
MT1A
TIPARP
UPP2
ACTA2
CLDN23
C3orf52
PLEKHA4
RAB39A
IL11
RALGDS
CACNG6
RGS22
PDK4
FAM134B
SPATA13
PSME1
RARRES3
PSMB10
CASP7
DUSP5
ST3GAL4
LDLR
RNF144A
B4GALT5
CXCL1
UBD
FZD5
BCL3
CEACAM1
TRIM21
OAS1
MYD88
OPTN
STAT1
CCND3
BST2
GBP3
UBE2L6
SP100
GBP4
CD68
CNP
PSMB9
APOL2
HLA−E
TAP1
IRF1
PSMB8
MDK
LY6E
TYMP
HLA−F
PMAIP1
PSME2
TRAFD1
NAPA
PDGFRL
HES4
ANGPTL4
RND1
NFKBIZ
CASP4
CFB
F2RL1
EMP1
NRN1
IER5
TRIM31
PTGS2
NFE2L3
C1S
SUSD3
H1F0
TRIM38
NCOA7
REC8
MASTL
IGFBP1
DNAJC6
ZDHHC11
GOLGA8B
FAM46C
DKK3
MT1G
FLRT3
NRARP
STC1
NFIL3
NBPF10
RAB27A
SMOX
FNDC4
BCL6
KLF9
SOX21

### EDF4

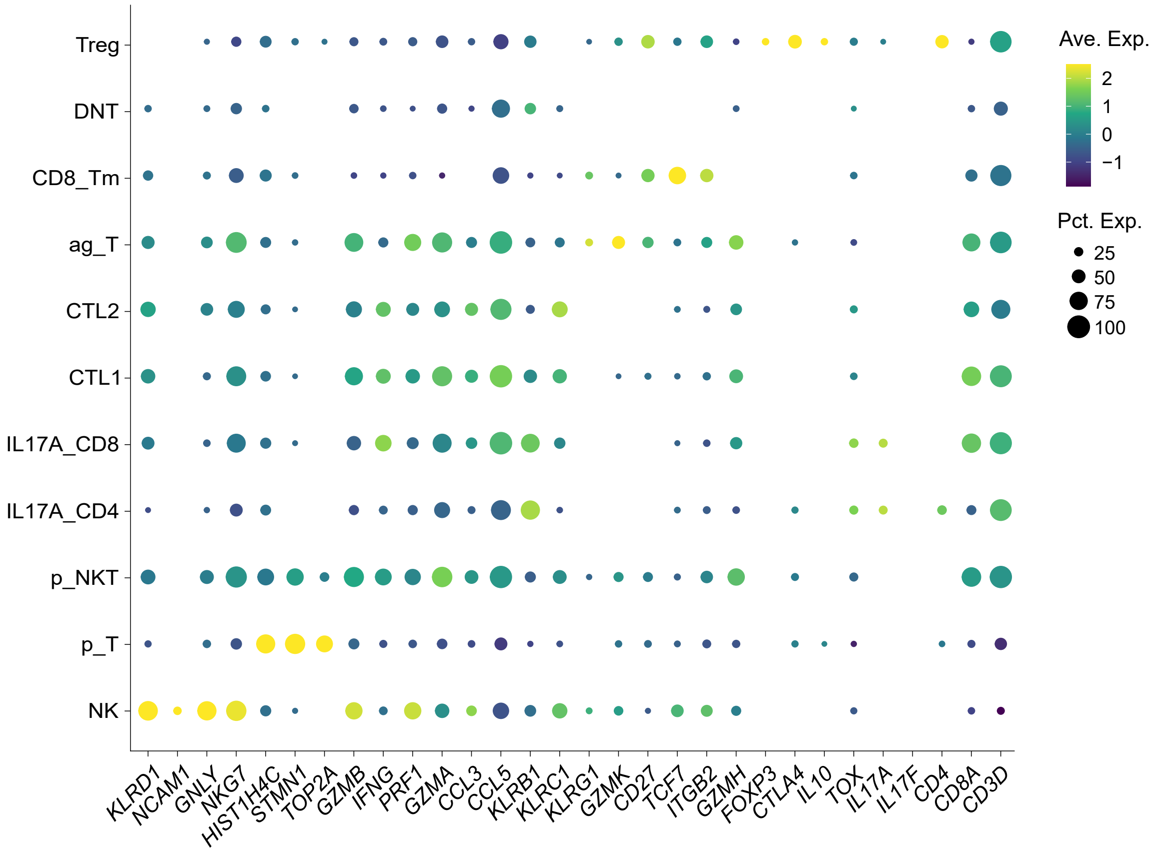

Treg
DNT
CD8_Tm
ag_T
CTL2
CTL1
IL17A_CD8
IL17A_CD4
p_NKT
p_T
NK
KLRD1
NCAM1
GNLY
NKG7
HIST1H4C
STMN1
TOP2A
GZMB
IFNG
PRF1
GZMA
CCL3
CCL5
KLRB1
KLRC1
KLRG1
GZMK
CD27
TCF7
ITGB2
GZMH
FOXP3
CTLA4
IL10
TOX
IL17A
IL17F
CD4
CD8A
CD3D
Ave. Exp.
2
1
0
−1
Pct. Exp.
25
50
75
100

**Supplementary Table 1:** Characteristics of the **(a)** positive (n = 45) and **(b)** negative (n = 42) individuals included in the here present study.

**(a)**

| **Patient-ID** | **Gender** | **Age** (years) | **SARS-CoV-2** | **WHO** | **Sampling dps*** | **Sampling dpP#** |
|---|---|---|---|---|---|---|
| BIH-c-CoV-01 | f | < 1 year | positive | mild | 8 | 0 |
| BIH-c-CoV-02 | m | < 1 year | positive | mild | 3 | 3 |
| BIH-c-CoV-03 | m | 1-5 | positive | asymptomatic | no symptoms | 2 |
| BIH-c-CoV-04 | f | 6-10 | positive | mild | 1 | 1 |
| BIH-c-CoV-05 | m | 6-10 | positive | mild | 11 | 6 |
| BIH-c-CoV-06 | m | 6-10 | positive | mild | 1 | 0 |
| BIH-c-CoV-07 | f | 6-10 | positive | moderate | 7 | 0 |
| BIH-c-CoV-08 | m | 6-10 | positive | mild | 4 | 1 |
| BIH-c-CoV-09 | m | 6-10 | positive | mild | 6 | 0 |
| BIH-c-CoV-10 | f | 11-15 | positive | asymptomatic | no symptoms | 0 |
| BIH-c-CoV-11 | m | 11-15 | positive | mild | 2 | 0 |
| BIH-c-CoV-12 | m | 11-15 | positive | moderate | 9 | 0 |
| BIH-c-CoV-13 | m | 11-15 | positive | mild | 3 | 2 |
| BIH-c-CoV-14 | m | 16-20 | positive | moderate | 2 | 2 |
| BIH-c-CoV-15 | f | 16-20 | positive | mild | 5 | 4 |
| BIH-c-CoV-16 | f | 16-20 | positive | mild | 3 | 0 |
| BIH-c-CoV-17 | m | 1-5 | positive | moderate | 3 | 4 |
| BIH-c-CoV-18 | m | 1-5 | positive | moderate | 0 | 4 |
| BIH-c-CoV-19 | f | 1-5 | positive | mild | 6 | 3 |
| BIH-c-CoV-20 | f | 6-10 | positive | mild | 6 | 6 |
| BIH-c-CoV-21 | m | 6-10 | positive | mild | 5 | 0 |
| BIH-c-CoV-22 | f | 6-10 | positive | mild | 5 | 0 |
| BIH-c-CoV-23 | f | 11-15 | positive | moderate | 2 | 8 |
| BIH-c-CoV-24 | m | 11-15 | positive | moderate | 6 | 7 |
| BIH-a-CoV-01 | f | 26-30 | positive | moderate | 2 | 2 |
| BIH-a-CoV-02 | f | 26-30 | positive | mild | 6 | 0 |
| BIH-a-CoV-03 | f | 31-35 | positive | mild | 4 | 0 |
| BIH-a-CoV-04 | f | 31-35 | positive | mild | 8 | 0 |
| BIH-a-CoV-05 | f | 31-35 | positive | moderate | 5 | 3 |
| BIH-a-CoV-06 | m | 31-35 | positive | moderate | 0 | 0 |
| BIH-a-CoV-07 | f | 36-40 | positive | moderate | 12 | 0 |
| BIH-a-CoV-08 | f | 41-45 | positive | moderate | 10 | 7 |
| BIH-a-CoV-09 | f | 41-45 | positive | moderate | 4 | 6 |
| BIH-a-CoV-10 | m | 41-45 | positive | moderate | 11 | 2 |
| BIH-a-CoV-11 | m | 46-50 | positive | moderate | 11 | 0 |
| BIH-a-CoV-12 | f | 46-50 | positive | moderate | 10 | 3 |
| BIH-a-CoV-13 | m | 46-50 | positive | mild | 3 | 6 |
| BIH-a-CoV-14 | f | 31-35 | positive | mild | 2 | 0 |
| BIH-a-CoV-15 | m | 31-35 | positive | mild | 0 | 1 |
| BIH-a-CoV-16 | m | 31-35 | positive | mild | 0 | 0 |
| BIH-a-CoV-17 | f | 36-40 | positive | mild | 4 | 0 |
| BIH-a-CoV-18 | f | 36-40 | positive | mild | 2 | 0 |
| BIH-a-CoV-19 | m | 76-80 | positive | moderate | 3 | 1 |
| BIH-a-CoV-20 | m | 46-50 | positive | mild | 4 | 0 |
| BIH-a-CoV-21 | m | 41-45 | positive | moderate | 3 | 7 |

*days post onset of symptoms; #days post first positive PCR

**Supplementary Table 1:** Continued.

**(b)**

| Patient-ID | Gender | Age (years) | SARS-CoV-2 | WHO | Sampling dps* |
|---|---|---|---|---|---|
| BIH-c-Con-01 | f | 1-5 | negative | negative | no symptoms |
| BIH-c-Con-02 | m | 1-5 | negative | negative | no symptoms |
| BIH-c-Con-03 | f | 1-5 | negative | negative | no symptoms |
| BIH-c-Con-04 | f | 1-5 | negative | negative | no symptoms |
| BIH-c-Con-05 | m | 6-10 | negative | negative | no symptoms |
| BIH-c-Con-06 | f | 6-10 | negative | negative | no symptoms |
| BIH-c-Con-07 | m | 6-10 | negative | negative | no symptoms |
| BIH-c-Con-08 | m | 6-10 | negative | negative | no symptoms |
| BIH-c-Con-09 | m | 6-10 | negative | negative | no symptoms |
| BIH-c-Con-10 | f | 6-10 | negative | negative | no symptoms |
| BIH-c-Con-11 | f | 6-10 | negative | negative | no symptoms |
| BIH-c-Con-12 | m | 6-10 | negative | negative | no symptoms |
| BIH-c-Con-13 | f | 6-10 | negative | negative | no symptoms |
| BIH-c-Con-14 | m | 11-15 | negative | negative | no symptoms |
| BIH-c-Con-15 | m | 11-15 | negative | negative | no symptoms |
| BIH-c-Con-16 | f | 11-15 | negative | negative | no symptoms |
| BIH-c-Con-17 | m | 11-15 | negative | negative | no symptoms |
| BIH-c-Con-18 | m | 16-20 | negative | negative | no symptoms |
| BIH-a-Con-01 | m | 21-25 | negative | negative | no symptoms |
| BIH-a-Con-02 | m | 26-30 | negative | negative | no symptoms |
| BIH-a-Con-03 | f | 26-30 | negative | negative | no symptoms |
| BIH-a-Con-04 | m | 26-30 | negative | negative | no symptoms |
| BIH-a-Con-05 | f | 26-30 | negative | negative | no symptoms |
| BIH-a-Con-06 | m | 31-35 | negative | negative | no symptoms |
| BIH-a-Con-07 | f | 31-35 | negative | negative | no symptoms |
| BIH-a-Con-08 | f | 36-40 | negative | negative | no symptoms |
| BIH-a-Con-09 | m | 36-40 | negative | negative | no symptoms |
| BIH-a-Con-10 | f | 41-45 | negative | negative | no symptoms |
| BIH-a-Con-11 | f | 41-45 | negative | negative | no symptoms |
| BIH-a-Con-12 | m | 46-50 | negative | negative | no symptoms |
| BIH-a-Con-13 | f | 46-50 | negative | negative | no symptoms |
| BIH-a-Con-14 | f | 46-50 | negative | negative | no symptoms |
| BIH-a-Con-15 | f | 61-65 | negative | negative | no symptoms |
| BIH-a-Con-16 | f | 61-65 | negative | negative | no symptoms |
| BIH-a-Con-17 | m | 61-65 | negative | negative | no symptoms |
| BIH-a-Con-18 | f | 66-70 | negative | negative | no symptoms |
| BIH-a-Con-19 | f | 66-70 | negative | negative | no symptoms |
| BIH-a-Con-20 | m | 61-65 | negative | negative | no symptoms |
| BIH-a-Con-21 | f | 61-65 | negative | negative | no symptoms |
| BIH-a-Con-22 | m | 76-80 | negative | negative | no symptoms |
| BIH-a-Con-23 | m | 55-60 | negative | negative | no symptoms |

*days post onset of symptoms

**Supplementary Table 2:** Summary of cohort characteristics.

| | SARS-CoV-2 negative | SARS-CoV-2 positive |
|---|---|---|
| **children** | | |
| female | 8 | 10 |
| male | 10 | 14 |
| 0-6 years | 4 | 7 |
| 7-10 years | 9 | 7 |
| 11-18 years | 5 | 10 |
| median age (years) | 9.0 ± 3.8 | 9.0 ± 5.6 |
| mean dps* (days) | - | 4.5 ± 2.8 |
| mean dpP# (days) | - | 2.2 ± 2.5 |
| **adults** | | |
| female | 13 | 12 |
| male | 10 | 9 |
| median age (years) | 46.0 ± 16.3 | 39.0 ±10.4 |
| mean dps* (days) | - | 5.0 ± 3.7 |
| mean dpP# (days) | - | 1.8 ± 2.5 |

*days post onset of symptoms, #days post first positive PCR test
